# Disparities in COVID-19 Fatalities among Working Californians

**DOI:** 10.1101/2021.11.10.21266195

**Authors:** Kristin J. Cummings, John Beckman, Matthew Frederick, Robert Harrison, Alyssa Nguyen, Robert Snyder, Elena Chan, Kathryn Gibb, Andrea Rodriguez, Jessie Wong, Erin L. Murray, Seema Jain, Ximena Vergara

## Abstract

**Background:** Information on the occupational distribution of COVID-19 mortality is limited.

**Objective:** To characterize COVID-19 fatalities among working Californians.

**Design:** Retrospective study of laboratory-confirmed COVID-19 fatalities with dates of death from January 1 to December 31, 2020.

**Setting:** California.

**Participants:** COVID-19 accounted for 8,050 (9.9%) of 81,468 fatalities among Californians 18-64 years old. Of these decedents, 2,486 (30.9%) were matched to state employment records and classified as “confirmed working.” The remainder were classified as “likely working” (n=4,121 [51.2%]) or “not working” (n=1,443 [17.9%]) using death certificate and case registry data.

**Measurements:** We calculated age-adjusted overall and occupation-specific COVID-19 mortality rates using 2019 American Community Survey denominators.

**Results:** Confirmed and likely working COVID-19 decedents were predominantly male (76.3%), Latino (68.7%), and foreign-born (59.6%), with high school or less education (67.9%); 7.8% were Black. The overall age-adjusted COVID-19 mortality rate was 30.0 per 100,000 workers (95% confidence interval [CI], 29.3-30.8). Workers in nine occupational groups had mortality rates higher than this overall rate, including those in farming (78.0; 95% CI, 68.7-88.2); material moving (77.8; 95% CI, 70.2-85.9); construction (62.4; 95% CI, 57.7-67.4); production (60.2; 95% CI, 55.7-65.0); and transportation (57.2; 95% CI, 52.2-62.5) occupations. While occupational differences in mortality were evident across demographic groups, mortality rates were three-fold higher for male compared with female workers and three- to seven-fold higher for Latino and Black workers compared with Asian and White workers.

**Limitations:** The requirement that fatalities be laboratory-confirmed and the use of 2019 denominator data may underestimate the occupational burden of COVID-19 mortality.

**Conclusion:** Californians in manual labor and in-person service occupations experienced disproportionate COVID-19 mortality, with the highest rates observed among male, Latino, and Black workers.

## Introduction

Following the identification of community transmission of SARS-CoV-2 in California in late February 2020, the state became the first in the nation to issue a stay-at-home order on March 19, 2020 (1-4). The order identified critical infrastructure sectors, including health, emergency services, food and agriculture, and transportation and logistics, in which Californians could work outside of the home (4, 5). As a result, at least 4.7 million Californians, or 25% of the entire workforce, continued to work in person with coworkers and/or members of the public (6).

Working-age adults (18-64 years old) account for 75% of confirmed COVID-19 cases and 29% of confirmed COVID-19 deaths in California (7). Yet, aside from descriptions of workplace cases and outbreaks (3, 8-10), little is known about the occupational distribution of COVID-19 morbidity and mortality in the state. A study of deaths that occurred from March to November 2020 among Californians 18-65 years old found relative excess all-cause mortality in specific occupational sectors, suggesting differential impact of COVID-19 on California’s workers (11). To better understand these dynamics and inform preventive interventions, we aimed to characterize COVID-19 fatalities that occurred in 2020 among working Californians, describe the distribution of occupations, and examine temporal trends.

## Methods

### Data Sources

We used three data sources to identify working Californians who died of COVID-19 in 2020. These were: 1) the Electronic Death Registration System (EDRS), which contains California’s death certificates; 2) the state’s COVID-19 Case Registry, comprising all laboratory-confirmed COVID-19 cases among California residents reported to the California Department of Public Health (CDPH) by local health jurisdictions; and 3) Employment Development Department (EDD) records derived from quarterly tax reports submitted to the EDD by employers subject to the state’s Unemployment Insurance laws and by federal agencies in California subject to the Unemployment Compensation for Federal Employees program (12).

We used local health jurisdiction determinations included in the COVID-19 Case Registry as of April 15, 2021 to define COVID-19 decedents; all other deaths were considered non-COVID decedents. Case Registry records of COVID-19 decedents were probabilistically matched to an EDRS dynamic file dated April 21, 2021 using first, middle, and last names; date of birth; date of death; zip code; city; county; and cause of death. The final dataset was restricted to decedents who were 18-64 years old at death. Age and other demographic characteristics were derived from the EDRS records.

We used EDD records to examine decedents’ 2020 employment by calendar quarter. Exact matching between EDRS and EDD records used social security number (SSN) from the death certificates. Members of the armed forces, the self-employed, proprietors, domestic workers, unpaid family workers, and railroad workers are not covered by the unemployment insurance systems noted above and are therefore excluded from the EDD data (12). In addition, workers in the “underground economy,” defined by EDD as “individuals and businesses that deal in cash and/or use other schemes to conceal their activities…from government” are not captured by EDD data (13).

### Working Status Classification

We classified decedents as *confirmed working* at the time of infection if EDD records indicated that the decedent was employed during the quarter of death or the previous quarter. Decedents were classified as *not working* at the time of infection and excluded if the EDRS or COVID-19 Case Registry records indicated that the decedent was unemployed, retired, incarcerated, not paid, a student, or a homemaker. Because EDD records cover only a subset of the workforce, a decedent who did not meet criteria for *confirmed working* or *not working* was classified as *likely working* at the time of infection and included.

### Statistical Analyses

U.S. Census Occupation Codes (2010) were assigned to decedents’ death certificate free text for “usual occupation” using a machine learning-based system, the National Institute for Occupational Safety and Health (NIOSH) Industry and Occupation Computerized Coding System (NIOCCS) (14). We conducted manual coding when NIOCCS reported less than 90% confidence of accuracy for an autocode.

The U.S. Census Bureau’s 2019 American Community Survey (ACS) Public Use Microdata Sample files for California were extracted by detailed occupation, age, sex, and race/ethnicity (15). We calculated crude overall, occupational group-specific, and detailed occupation-specific COVID-19 mortality rates by combining confirmed and likely working decedents for numerators and using ACS data to generate denominators. Rates were calculated for five or more deaths per occupational group, detailed occupation, or by subgroup. Deaths were aggregated at the occupational group level: 1) overall, 2) by sex, and 3) by race/ethnicity to calculate age-adjusted rates. Direct age standardization to the ACS employment data was performed using five age groups: 18-29, 30-39, 40-49, 50-59, and 60-64 years. Combined age-, sex- and race/ethnicity-standardization was not possible due to sparse data.

We calculated a Prevention Index for each detailed occupation by averaging the rank orders of fatality counts and crude mortality rates (16).

Statistical analyses were conducted using SAS software V.9.4 (SAS Institute, Inc, Cary, North Carolina, USA) and R Studio Version 4.0.2 (R Studio, PBC, Boston, Massachusetts, USA). Rates were calculated using the R epitools package.

### Ethical Considerations

The California Health and Human Services Agency’s Committee for the Protection of Human Subjects exempted the study from institutional review board review.

## Results

### 2020 California Decedents

Among 317,894 people who died in California in 2020, we identified 81,468 (25.6%) who were 18-64 years old at the time of death, including 8,050 (9.9%) COVID-19 decedents (Figure 1). Among the COVID-19 decedents, 2,486 (30.9%) were classified as confirmed working, 4,121 (51.2%) as likely working, and 1,443 (17.9%) as not working. The number of COVID-19 decedents by day reflected the state’s overall epidemic curve,^7^ with the proportions by working status relatively stable throughout 2020 (Figure 2).

**Figure 1.**
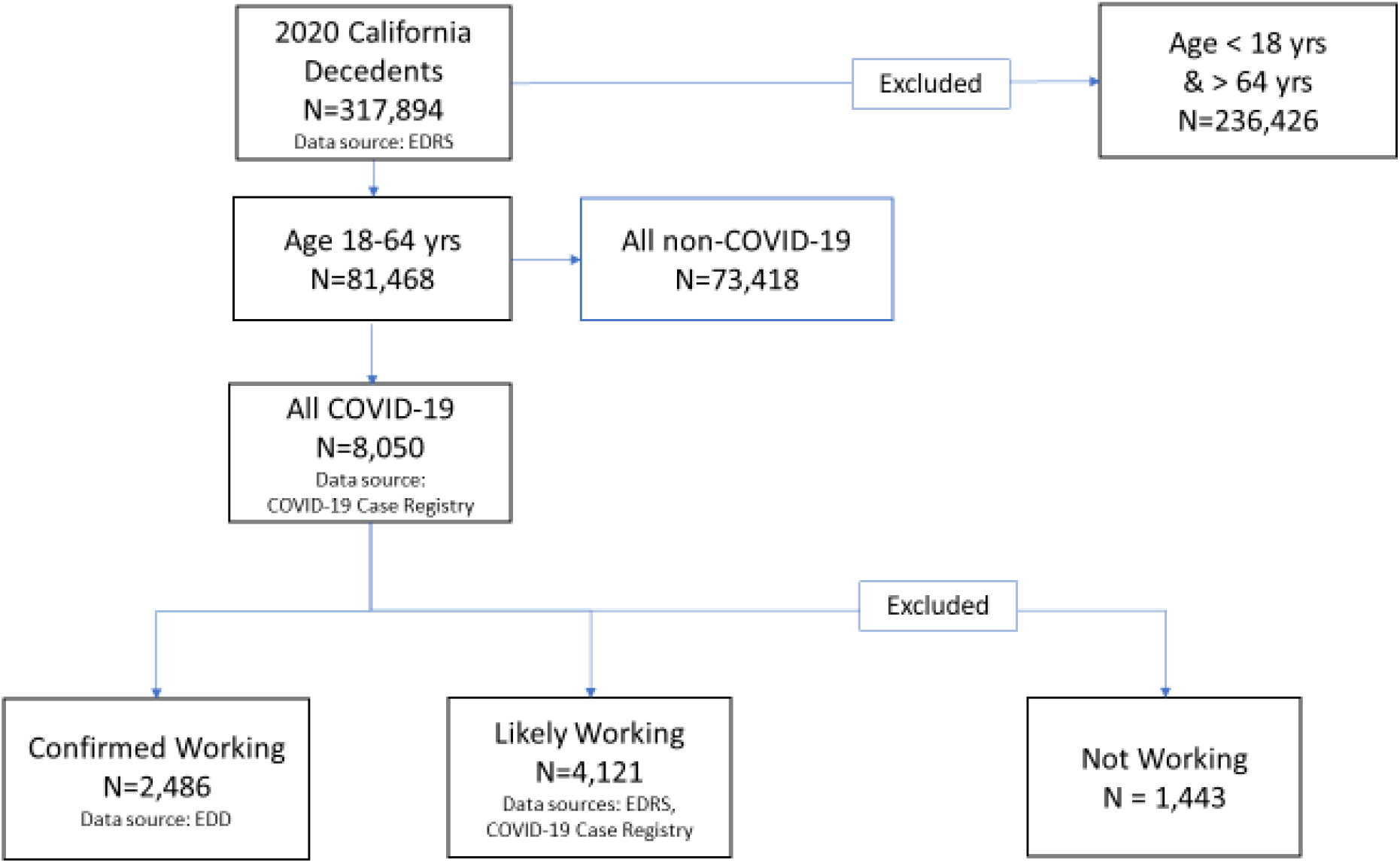
Identification of California’s 2020 COVID-19 Decedents Ages 18-64 Years and Classification According to Working Status. Laboratory-confirmed fatal COVID-19 cases were reported by local health jurisdictions to the California Department of Public Health. Working status was classified according to state employment records and information available on death certificates and in California’s COVID-19 Case Registry. Abbreviations: EDRS, Electronic Death Registration System; EDD, Employment Development Department.

**Figure 2.**
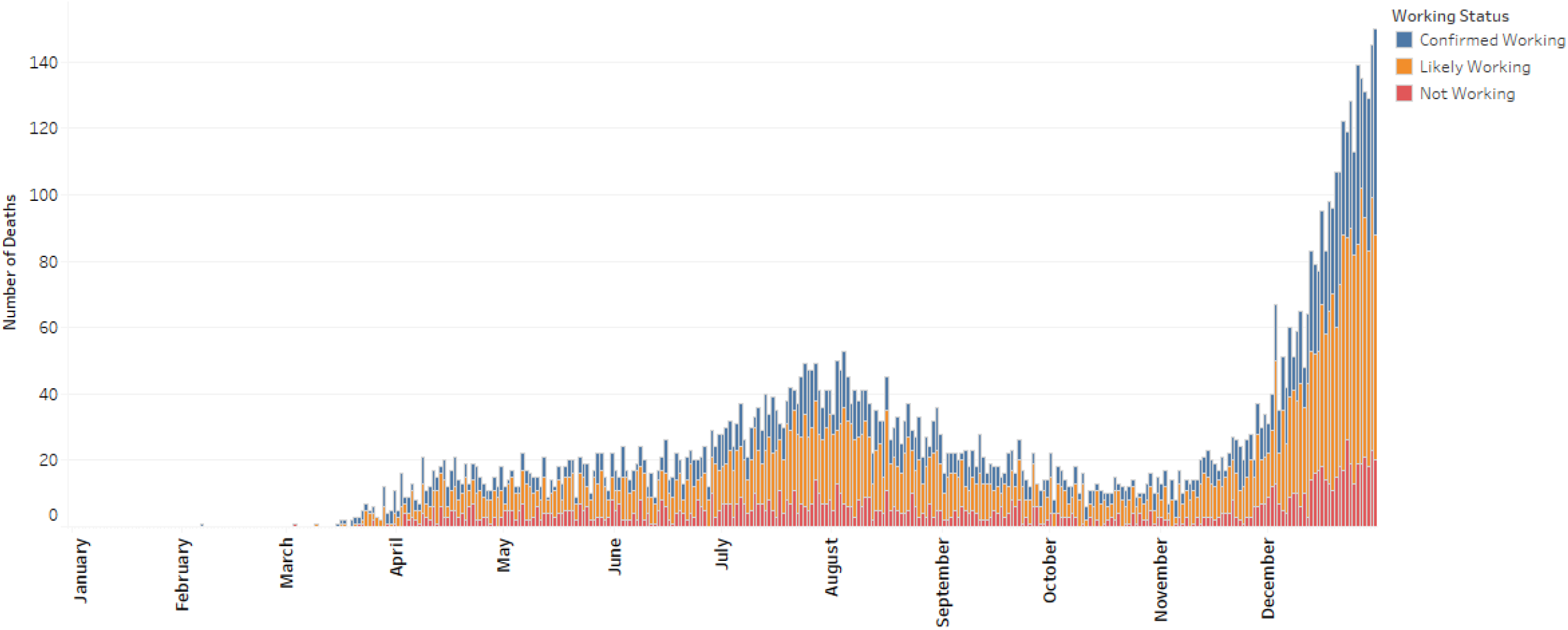
COVID-19 Deaths among Decedents Ages 18-64 Years, by Day and Decedents’ Working Status, California, 2020. Laboratory-confirmed fatal COVID-19 cases were reported by local health jurisdictions to the California Department of Public Health. Working status was classified according to state employment records and information available on death certificates and in California’s COVID-19 Case Registry.

Compared to non-COVID-19 decedents, all COVID-19 decedents, regardless of working status, more often were older, male, Latino, and foreign-born and had lower education levels (Table 1). A total of 7,214 (9.8%) non-COVID-19 decedents and 1,536 (19.1%) COVID-19 decedents lacked a valid social security number on the death certificate, including 1,222 (29.7%) likely working COVID-19 decedents. Compared to not working COVID-19 decedents, confirmed and likely working COVID-19 decedents more often were male and foreign-born and had higher education levels (Table 1). Compared to likely working decedents, confirmed working decedents more often were Asian and had higher education levels.

**Table 1.**
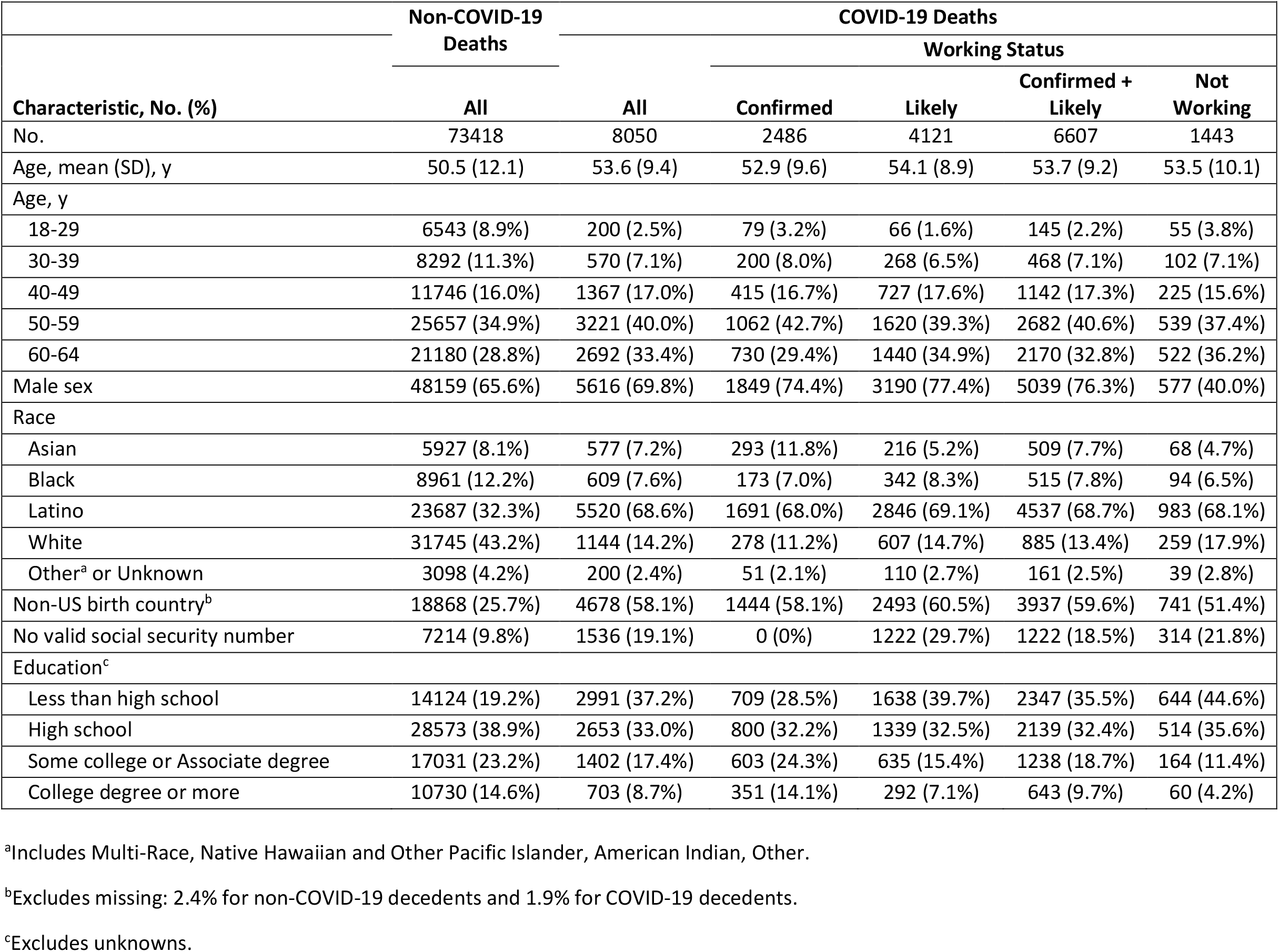
Non-COVID-19 Fatalities and COVID-19 Fatalities Overall and by Work Status among Decedents Ages 18-64 Years, California, January 1, 2020 - December 31, 2020.

### COVID-19 Mortality Rates by Occupational Group

The overall age-adjusted COVID-19 mortality rate was 30.0 deaths per 100,000 workers (95% CI, 29.3-30.8) (Figure 3A). Nine occupational groups had mortality rates higher than this overall rate: farming, fishing, and forestry (78.0 per 100,000; 95% CI, 68.7-88.2); material moving (77.8; 95% CI, 70.2-85.9); construction and extraction (62.4; 95% CI, 57.7-67.4); production (60.2; 95% CI, 55.7-65.0); transportation (57.2; 95% CI, 52.2-62.5); installation, maintenance, and repair (55.2; 95% CI, 49.3-61.7); building and grounds cleaning and maintenance (46.9; 95% CI, 42.7-51.5); food preparation and serving related (46.0; 95% CI, 41.2-51.1); and protective service (44.0; 95% CI, 37.8-50.9). The proportion of COVID-19 fatalities by occupational group did not vary substantially over time (data not shown).

**Figure 3.**
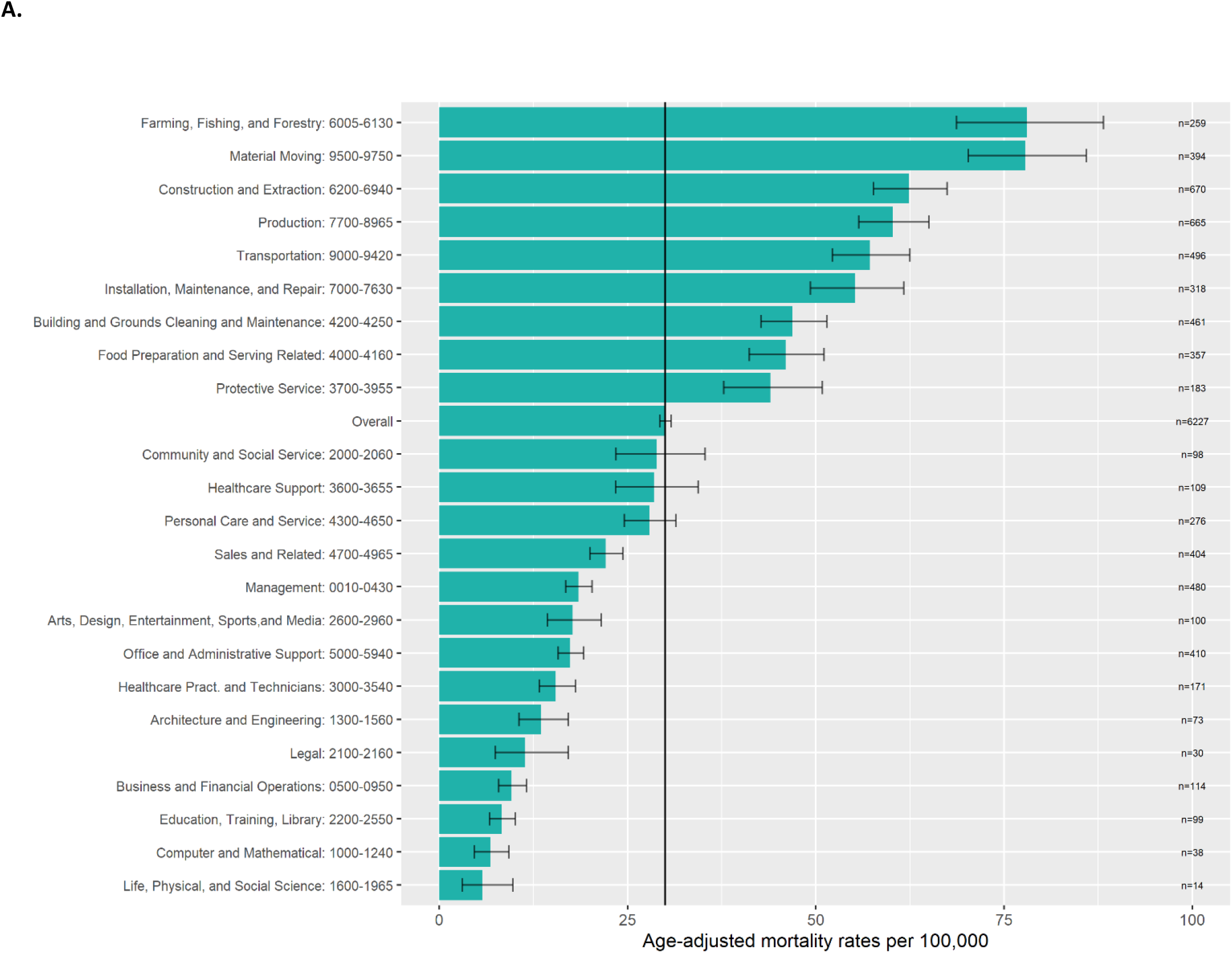
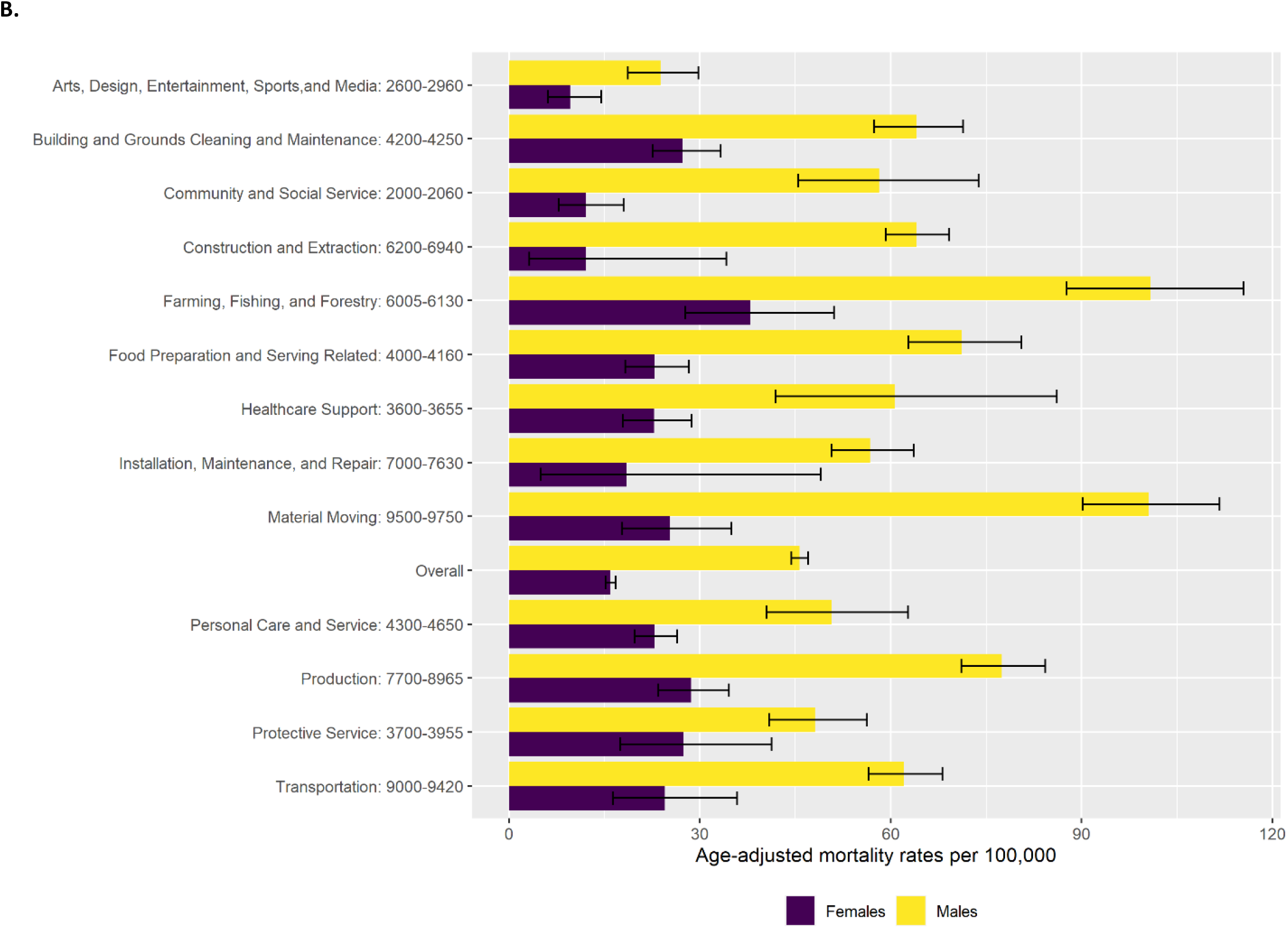
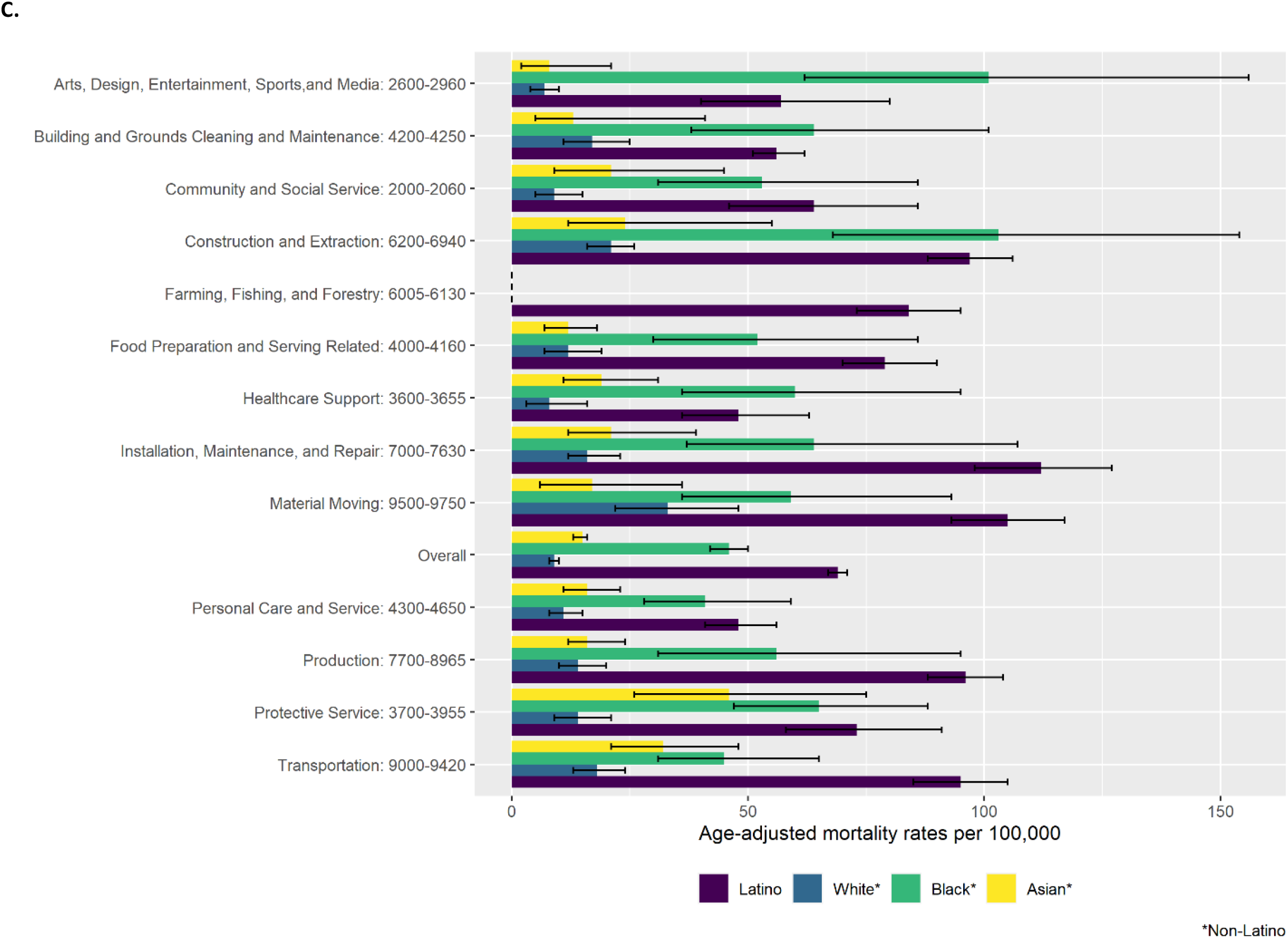
Age-adjusted COVID-19 Mortality Rates among Workers Ages 18-64 Years, California, 2020. (A) Overall and for all occupational groups. (B) Overall and for select occupational groups, by sex. (C) Overall and for select occupational groups, by race/ethnicity. Information on occupations included within each occupational group is available from the U.S. Census Bureau (https://www.bls.gov/cps/cenocc2010.htm).

The overall age-adjusted COVID-19 mortality rate for male workers (45.7 per 100,000; 95% CI, 44.4-47.0) was nearly three-fold higher than for female workers (15.9; 95% CI, 15.2-16.8) (Figure 3B). This pattern was evident for occupation-specific rates as well. Nonetheless, higher-mortality occupational groups were evident for female workers. For instance, female workers in farming, fishing, and forestry had a mortality rate (38.0; 95% CI 27.7-51.1) that was more than twice the rate of female workers overall.

Overall age-adjusted COVID-19 mortality rates were three- to seven-fold higher for Latino (69.1 per 100,000; 95% CI 67.1.-71.2) and Black (46.4; 95% CI, 42.5-50.6) workers than for Asian (15.0; 95% CI, 13.7-16.4) and White (9.5; 95% CI, 8.9-10.2) workers; similar patterns were observed for occupation-specific rates (Figure 3C). Mortality rates were highest for Latino workers in installation, maintenance, and repair (112.1 per 100,000; 95% CI, 98.2-127.4); material moving (105.1; 95% CI, 93.8-117.4); and construction (97.1; 95% CI 88.4-106.6) occupations, and for Black workers in construction and extraction (103.9; 95% CI, 68.1-154.6); arts, design, entertainment, sports, and media (101.2; 95% CI, 62.6-154.6), and protective services (65.4; 95% CI, 47.2-88.6) occupations. Despite lower mortality rates among Asian and White workers compared to Latino and Black workers, higher-mortality occupational groups were evident for Asian and White workers. For instance, Asian workers in protective services occupations (46.5; 95% CI, 26.5-75.9) had a mortality rate that was three-fold higher than the rate for Asian workers overall; the rate for White workers in material moving occupations (33.6; 95% CI, 22.6-48.5) was more than three times higher than the rate for White workers overall.

Age-adjusted COVID-19 mortality rates for healthcare occupational groups were not elevated overall (Figure 3A). For healthcare support occupations, male (60.7; 95% CI, 41.9-86.1), Black (60.4; 95% CI, 36.2-95.6), and Latino (48.2; 95% CI, 36.4-63.1) workers had mortality rates higher than the overall rate (28.5; 95% CI, 23.4-34.4); female (22.8; 95% CI, 17.9-28.7), Asian (19.6; 95% CI, 11.5-31.6), and White (8.4; 95% CI, 3.8-16.6) workers had lower rates (Figure 3B-C). For healthcare practitioner and technician occupations, male (23.3; 95% CI, 18.3-29.4), Latino (35.6; 95% CI, 26.4-47.3), Black (24.4; 95% CI, 14.4-41.6), and Asian (21.0; 95% CI, 16.1-27.3) workers had mortality rates higher than the overall rate (15.5; 95% CI, 13.3-18.1); female (12.2; 95% CI, 9.9-15.0) and White (5.2; 95% CI, 3.5-7.9) workers had lower rates.

### Detailed Occupations

The detailed occupation that ranked highest on the Prevention Index was sewing machine operators, followed by construction laborers and automotive service technicians and mechanics (Table 2). The ratio of likely working to confirmed working status among decedents in the top 20 detailed occupations ranged from 0.5 for bus drivers to 6.4 for sewing machine operators, with an average of 2.5.

**Table 2.** Top 20 Detailed Occupations Ranked by Mortality Prevention Index among Workers Ages 18-64 Years, California, January 1, 2020 - December 31, 2020.

|  |  |  |  |  |  |  |  |
| --- | --- | --- | --- | --- | --- | --- | --- |
| **Detailed Occupation (2010 Census Code)** | **Deaths by Working Status, No.** |  |  | **Employment** | **Crude Rate<sup>a</sup>** | **PI Rank** | **Likely: Confirmed<sup>b</sup>** |
|  | **Confirmed** | **Likely** | **Total** |  |  |  |  |
| Sewing machine operators (8320) | 11 | 70 | 81 | 34906 | 232 | 1 | 6.4 |
| Construction laborers (6260) | 66 | 277 | 343 | 331577 | 103 | 2 | 4.2 |
| Automotive service technicians and mechanics (7200) | 24 | 92 | 116 | 110061 | 105 | 3 | 3.8 |
| Driver/sales workers and truck drivers (9130) | 139 | 227 | 366 | 439063 | 83 | 4 | 1.6 |
| Farmers, ranchers, and other agricultural managers (0205) | 21 | 29 | 50 | 40803 | 123 | 5 | 1.4 |
| Chefs and head cooks (4000) | 21 | 60 | 81 | 84574 | 96 | 6 | 2.9 |
| Industrial truck and tractor operators (9600) | 40 | 41 | 81 | 83574 | 97 | 7 | 1.0 |
| Miscellaneous agricultural workers (6050) | 82 | 145 | 227 | 296595 | 77 | 8 | 1.8 |
| Laborers and freight, stock, and material movers, hand (9620) | 99 | 147 | 246 | 335476 | 73 | 9 | 1.5 |
| Security guards and gaming surveillance officers (3930) | 58 | 73 | 131 | 167976 | 78 | 10 | 1.3 |
| Machinists (8030) | 16 | 25 | 41 | 36466 | 112 | 11 | 1.6 |
| Clergy (2040) | 5 | 31 | 36 | 31718 | 114 | 12 | 6.2 |
| Grounds maintenance workers (4250) | 33 | 104 | 137 | 199603 | 69 | 13 | 3.2 |
| Janitors and building cleaners (4220) | 84 | 102 | 186 | 300067 | 62 | 14 | 1.2 |
| Welding, soldering, and brazing workers (8140) | 11 | 35 | 46 | 57444 | 80 | 15 | 3.2 |
| First-line supervisors of housekeeping and janitorial workers (4200) | 12 | 18 | 30 | 27015 | 111 | 16 | 1.5 |
| Supervisors of transportation and material moving workers (9000) | 19 | 12 | 31 | 29719 | 104 | 17 | 0.6 |
| Bakers (7800) | 10 | 24 | 34 | 37374 | 91 | 18 | 2.4 |
| Painting workers (8810) | 5 | 18 | 23 | 19675 | 117 | 19 | 3.6 |
| Bus drivers (9120) | 26 | 13 | 39 | 52137 | 75 | 20 | 0.5 |

Abbreviation: PI, prevention index
<sup>a</sup>Crude rate per 100,000 workers.
<sup>b</sup>Ratio of likely working to confirmed working decedents within detailed occupation.

## Discussion

We found that COVID-19 accounted for 10% of all deaths among working-age Californians in 2020. This burden was unevenly distributed across the state’s population. Among those COVID-19 decedents identified as confirmed or likely working at the time of infection, most were male, Latino, and foreign-born, with a high school education or less. The highest age-adjusted COVID-19 mortality rates were in occupational groups in which work is typically carried out in person: farming, material moving, construction, production, transportation, installation, cleaning, and food service.

These findings are consistent with prior studies of COVID-19 mortality among Californians and other populations. A study of excess all-cause mortality among Californians 18-65 years old during the first nine months of the pandemic found the highest relative and per capita excess mortality in food/agriculture, transportation/logistics, manufacturing, and facilities sectors (11). Investigation of COVID-19 deaths among people 16-64 years old in Massachusetts from March through July 2020 also documented above-average mortality rates in the occupational groups we identified, except farming (17). An occupational analysis of 2020 COVID-19 decedents aged 20-64 years in England and Wales found the highest mortality rates in those working in process plants, security, caring personal services, food service, and transportation (18).

We observed that age-adjusted COVID-19 mortality rates were highest for male, Latino, and Black workers, both overall and within specific occupational groups. Similar patterns were noted in the study of COVID-19 mortality in Massachusetts (17). Our findings reflect a growing body of literature documenting a disproportionate burden of severe and fatal COVID-19 cases among men and racial/ethnic minorities (19-25). It is important to note that the occupational disparities in COVID-19 fatalities that we found could not be attributed solely to differences in employment by sex or race/ethnicity. Within each demographic subset, there was a gradient of occupational risk, with remarkable consistency in terms of which occupational groups had higher mortality. One exception was the high mortality rate among Black decedents in arts, design, entertainment, sports, and media occupations. These findings, coupled with the temporal consistencies in working status and occupation we observed, suggest specific occupational risks for California’s workers and highlight the intersecting contributions of occupation and other socioeconomic factors to COVID-19 health inequities (26-28).

In contrast to Massachusetts, England, and Wales (17, 18), we did not find above-average COVID-19 mortality in healthcare occupations in California. In Massachusetts, healthcare support occupations had the highest age-adjusted mortality rate of all occupational groups at nearly three times the average (17). In England and Wales, care workers, home carers, nursing auxiliaries and assistants, and nurses were among the occupations with the highest mortality rates (18). In California, we found that male, Latino, Black, and, in some cases, Asian workers in healthcare occupations had mortality rates that were higher than the overall rates for these occupational groups, but healthcare occupations were not identified for prioritization by the Prevention Index. The reasons for these discrepancies are uncertain, but they could reflect differences in the time course of the pandemic and COVID-19 deaths. Massachusetts and the United Kingdom were impacted early, with peak numbers of daily deaths for 2020 occurring on days in April (29, 30), whereas deaths peaked in California in December (Figure 2). Given the improvements in understanding of disease transmission and access to personal protective equipment that occurred over the course of 2020 (31-36), California may have been better positioned to protect its healthcare workers during the state’s later epidemic peak. In addition, the potential contribution of California’s longstanding Aerosol Transmissible Diseases standard, which requires healthcare employers to have written safety plans, provide personal protective equipment including respirators for protection from novel pathogens (considered under the standard to be potentially airborne), and train employees on safety procedures, merits further inquiry (37).

A unique strength of our study is the use of state employment records to confirm working status of COVID-19 decedents. Death certificates collect information about a decedent’s “usual” occupation, but do not confirm that the decedent was employed at the time of death (38). Yet the increase in specificity imparted by state employment records comes at a cost of decreased sensitivity, as many workers are excluded from official employment statistics (12). In particular, we were concerned about the exclusion of workers in the underground economy, such as day laborers and independent contractors, who make up substantial proportions of some California industries and may be more vulnerable on account of socioeconomic disadvantages (39-41). Thus, we included a “likely working” group of decedents who did not match to state employment records but for whom there was no documentation of not working status. Over 60% of the likely working decedents were foreign-born and nearly a third lacked a valid social security number, suggesting this group included undocumented immigrants who continued to work in person out of economic necessity (42). Notably, among the top 20 detailed occupations identified by the Prevention Index, the ratio of likely working to confirmed working decedents (2.5) exceeded the ratio for all 6,607 working decedents (1.7).

Despite our use of state employment records to confirm employment, our analyses were not designed to assess a causal relationship between employment and COVID-19 infection and death. The COVID-19 deaths that we identified among working decedents may have resulted from transmission of infection in the workplace, at home, or elsewhere in the community. Yet, there is reason to think that work contributed to the high COVID-19 mortality rates in certain occupational groups. The occupations with highest mortality in California overlap with the critical infrastructure sectors that were exempted from that state’s stay-at-home order (5). These occupations tend to involve manual labor or in-person provision of services that cannot be done remotely, and many require proximity to others at work (43, 44). Thus, the conditions of work in these high-mortality occupations may have created opportunities for workplace exposure to SARS-CoV-2. Workplace outbreaks provide further evidence. In Los Angeles County, nearly 60% of non-residential, non-healthcare workplace outbreaks that occurred through September 30, 2020 were in industrial sectors related to many of the occupations that we identified as having elevated mortality: manufacturing, retail trade, and transportation and warehousing (10). In Toronto, Canada, a similar analysis through June 2020 found the majority (68%) of workplace outbreaks in manufacturing, agriculture, and transportation and warehousing (45). In Europe, analysis of outbreak data from March through July 2020 revealed large numbers of clusters in the food packaging and processing sectors, in factories and manufacturing, and in office settings (46).

This study has several limitations. Our definition of COVID-19 fatality required a positive polymerase chain reaction (PCR) test result for SARS-CoV-2. Thus, COVID-19 decedents in California who did not undergo PCR testing were excluded, potentially underestimating the occupational burden of COVID-19 deaths. The working status of some decedents may have been misclassified, such as unemployed decedents classified as likely working. Our use of “usual occupation” listed on the death certificate may have misclassified the occupation of some decedents who were working in a different occupation at the time of infection. In addition, we used ACS data from 2019, as 2020 data were not yet available. Pandemic-related changes in employment that occurred in California in 2020 may mean that denominators were overestimated (47). As a result, we may have underestimated the COVID-19 mortality rates for some occupations. Finally, by focusing on working-age decedents, we did not address the potential burden of COVID-19 on workers 65 years of age and older.

Our findings have implications for prevention. Given the likelihood that COVID-19 fatalities among working Californians included work-related cases, the occupations with elevated mortality rates should be prioritized by public health and regulatory authorities to ensure that non-pharmaceutical interventions, such as physical distancing, use of respiratory protection or face coverings, and adequate ventilation, are implemented in the workplace (37, 48, 49). These high-mortality occupations also should be highlighted in COVID-19 vaccination campaigns, as they represent populations at high risk of severe outcomes regardless of where transmission occurs. As we plan for future respiratory viral pandemics, the occupational burden of COVID-19 mortality in California needs to be considered when crafting policies to mitigate disease transmission, including early implementation of non-pharmaceutical interventions in high-mortality occupations, rapid deployment of wage replacement programs that would allow those at high risk of poor outcomes to avoid in-person work, and prioritization of workers in high-mortality occupations for testing and vaccination when available (50, 51). More broadly, future pandemic responses must consider the concentration of historically disadvantaged racial/ethnic minorities, including undocumented immigrants, in high mortality occupations.

## Data Availability

Requests for data should be communicated to the California Department of Public Health.

## References

1. Zwald ML, Lin W, Sondermeyer Cooksey GL, et al. Rapid sentinel surveillance for COVID-19 — Santa Clara County, California, March 2020. MMWR Morb Mortal Wkly Rep. 2020;69:419–421.

2. Spellberg B, Haddix M, Lee R, et al. Community prevalence of SARS-CoV-2 among patients with influenza-like illnesses presenting to a Los Angeles Medical Center in March 2020. JAMA. 2020;323(19):1966–1967.

3. Heinzerling A, Stuckey MJ, Scheuer T, et al. Transmission of COVID-19 to health care personnel during exposures to a hospitalized patient -Solano County, California, February 2020. MMWR Morb Mortal Wkly Rep. 2020;69(15):472–476.

4. State of California. Executive Department. Executive Order N-33-20. Accessed November 8, 2021. https://www.gov.ca.gov/wp-content/uploads/2020/03/3.19.20-attested-EO-N-33-20-COVID-19-HEALTH-ORDER.pdf.

5. California State Public Health Officer. Essential workforce. Accessed November 8, 2021. https://covid19.ca.gov/img/EssentialCriticalInfrastructureWorkers.pdf.

6. State of California. Legislative Analyst’s Office. COVID-19 and the labor market: who are California’s frontline and remote workers? Accessed November 8, 2021. https://lao.ca.gov/LAOEconTax/Article/Detail/593.

7. State of California. COVID-19 cases dashboard. Accessed November 8, 2021. https://public.tableau.com/profile/ca.open.data#!/vizhome/COVID-19CasesDashboardv2_0/CaseStatistics.

8. Feaster M, Goh YY. High proportion of asymptomatic SARS-CoV-2 infections in 9 long-term care facilities, Pasadena, California, USA, April 2020. Emerg Infect Dis. 2020;26(10):2416–2419.

9. Waltenburg MA, Rose CE, Victoroff T, et al. Coronavirus Disease among workers in food processing, food manufacturing, and agriculture workplaces. Emerg Infect Dis. 2021;27(1):243–249.

10. Contreras Z, Ngo V, Pulido M, et al. Industry sectors highly affected by worksite outbreaks of coronavirus disease, Los Angeles County, California, USA, March 19-September 30, 2020. Emerg Infect Dis. 2021;27(7):1769–1775.

11. Chen YH, Glymour M, Riley A, et al. Excess mortality associated with the COVID-19 pandemic among Californians 18-65 years of age, by occupational sector and occupation: March through November 2020. PLoS One. 2021;16(6):e0252454.

12. State of California, Employment Development Department (EDD). Accessed November 8, 2021. Quarterly Census of Employment and Wages. https://www.labormarketinfo.edd.ca.gov/data/QCEW_About_the_Data.html.

13. State of California. Employment Development Department (EDD). Underground economy operations. Accessed November 8, 2021. https://edd.ca.gov/Payroll_Taxes/Underground_Economy_Operations.htm#DefinitionofUndergroundEconomy.

14. National Institute for Occupational Safety and Health. NIOSH Industry and Occupation Computerized Coding System (NIOCCS). Accessed November 8, 2021. https://csams.cdc.gov/nioccs/About.aspx.

15. U.S. Census Bureau. 2019 American Community Survey 1-year Public Use Microdata Samples [csv data file]. Accessed November 8, 2021. https://www.census.gov/programs-surveys/acs/microdata.html.

16. Bonauto D, Silverstein B, Adams D, Foley M. Prioritizing industries for occupational injury and illness prevention and research, Washington State workers’ compensation claims, 1999-2003. J Occup Environ Med. 2006;48(8):840–851.

17. Hawkins D, Davis L, Kriebel D. COVID-19 deaths by occupation, Massachusetts, March 1-July 31, 2020. Am J Ind Med. 2021;64(4):238–244.

18. United Kingdom Office of National Statistics. Coronavirus (COVID-19) related deaths by occupation, England and Wales: deaths registered between 9 March and 28 December 2020. Accessed November 8, 2021. https://www.ons.gov.uk/peoplepopulationandcommunity/healthandsocialcare/causesofdeath/bulletins/coronaviruscovid19relateddeathsbyoccupationenglandandwales/deathsregisteredbetween9marchand28december2020.

19. Gold JAW, Rossen LM, Ahmad FB, et al. Race, ethnicity, and age trends in persons who died from COVID-19 - United States, May-August 2020. MMWR Morb Mortal Wkly Rep. 2020;69(42):1517–1521.

20. Webb Hooper M, Nápoles AM, Pérez-Stable EJ. COVID-19 and racial/ethnic disparities. JAMA. 2020;323(24):2466–2467.

21. Rossen LM, Branum AM, Ahmad FB, Sutton P, Anderson RN. Excess deaths associated with COVID-19, by age and race and ethnicity - United States, January 26-October 3, 2020. MMWR Morb Mortal Wkly Rep. 2020;69(42):1522–1527.

22. Garcia E, Eckel SP, Chen Z, Li K, Gilliland FD. COVID-19 mortality in California based on death certificates: disproportionate impacts across racial/ethnic groups and nativity. Ann Epidemiol. 2021;58:69–75.

23. Gomez JMD, Du-Fay-de-Lavallaz JM, Fugar S, et al. Sex differences in COVID-19 hospitalization and mortality. J Womens Health (Larchmt). 2021;30(5):646–653.

24. Romano SD, Blackstock AJ, Taylor EV, et al. Trends in racial and ethnic disparities in COVID-19 hospitalizations, by region -United States, March-December 2020. MMWR Morb Mortal Wkly Rep. 2021;70(15):560–565.

25. Siegel M, Critchfield-Jain I, Boykin M, Owens A. Actual racial/ethnic disparities in COVID-19 mortality for the non-Hispanic Black compared to non-Hispanic White population in 35 US States and their association with structural racism. J Racial Ethn Health Disparities. 2021:1–13.

26. Bui DP, McCaffrey K, Friedrichs M, et al. Racial and ethnic disparities among COVID-19 cases in workplace outbreaks by industry sector - Utah, March 6-June 5, 2020. MMWR Morb Mortal Wkly Rep. 2020;69(33):1133–1138.

27. Mutambudzi M, Niedwiedz C, Macdonald EB, et al. Occupation and risk of severe COVID-19: prospective cohort study of 120 075 UK Biobank participants. Occup Environ Med. 2020. doi: 10.1136/oemed-2020-106731.

28. Côté D, Durant S, MacEachen E, et al. A rapid scoping review of COVID-19 and vulnerable workers: Intersecting occupational and public health issues. Am J Ind Med. 2021;64(7):551–566.

29. U.S. Department of Health and Human Services. COVID-19 state profile report – Massachusetts. Accessed November 8, 2021. https://healthdata.gov/Community/COVID-19-State-Profile-Report-Massachusetts/j75q-tgps.

30. Public Health England. Coronavirus (COVID-19) in the UK: Deaths in United Kingdom. Accessed November 8, 2021. https://coronavirus.data.gov.uk/details/deaths.

31. Prather KA, Marr LC, Schooley RT, McDiarmid MA, Wilson ME, Milton DK. Airborne transmission of SARS-CoV-2. Science. 2020;370(6514):303–304.

32. Tang JW, Bahnfleth WP, Bluyssen PM, et al. Dismantling myths on the airborne transmission of severe acute respiratory syndrome coronavirus-2 (SARS-CoV-2). J Hosp Infect. 2021;110:89–96.

33. Tang JW, Marr LC, Li Y, Dancer SJ. Covid-19 has redefined airborne transmission. BMJ. 2021;373:n913.

34. Centers for Disease Control and Prevention. Optimizing personal protective equipment (PPE) supplies. Accessed November 8, 2021. https://www.cdc.gov/coronavirus/2019-ncov/hcp/ppe-strategy/index.html.

35. Centers for Disease Control and Prevention. Scientific brief: SARS-CoV-2 transmission. Accessed November 8, 2021. https://www.cdc.gov/coronavirus/2019-ncov/science/science-briefs/sars-cov-2-transmission.html.

36. Food and Drug Administration. Update: FDA recommends transition from use of non-NIOSH-approved and decontaminated disposable respirators - letter to health care personnel and facilities. Accessed November 8, 2021. https://www.fda.gov/medical-devices/letters-health-care-providers/update-fda-recommends-transition-use-non-niosh-approved-and-decontaminated-disposable-respirators.

37. State of California. California Code of Regulations, Title 8, Section 5199. Aerosol Transmissible Diseases. Accessed November 8, 2021. https://www.dir.ca.gov/title8/5199.html.

38. Hawkins D. Differential occupational risk for COVID-19 and other infection exposure according to race and ethnicity. Am J Ind Med. 2020;63(9):817–820.

39. Graham MR, Ong P. 2007. Social, economic, spatial, and commuting patterns of informal jobholders. U.S. Census Bureau Technical Paper No. TP-2007-02. Accessed November 8, 2021. https://www2.census.gov/ces/tp/tp-2007-02.pdf.

40. Cummings KJ, Kreiss K. Contingent workers and contingent health: risks of a modern economy. JAMA. 2008; 299(4):448–450.

41. Yen Liu Y, Flaming D, Burns P. Sinking underground: the growing informal economy in California construction. Accessed November 8, 2021. https://economicrt.org/wp-content/uploads/2014/09/Sinking_Underground_2014.pdf.

42. Page KR, Flores-Miller A. Lessons we’ve learned - Covid-19 and the undocumented Latinx community. N Engl J Med. 2021;384(1):5–7.

43. Dingel JI, Neiman B. How many jobs can be done at home? J Public Econ. 2020;189:104235.

44. Cox-Ganser JM, Henneberger PK. Occupations by proximity and indoor/outdoor work: relevance to COVID-19 in all workers and black/hispanic workers. Am J Prev Med. 2021;60(5):621–628.

45. Murti M, Achonu C, Smith BT, et al. COVID-19 workplace outbreaks by industry sector and their associated household transmission, Ontario, Canada, January to June, 2020. J Occup Environ Med. 2021;63(7):574–580.

46. European Centre for Disease Prevention and Control. COVID-19 clusters and outbreaks in occupational settings in the EU/EEA and the UK. Accessed November 8, 2021. https://www.ecdc.europa.eu/en/publications-data/covid-19-clusters-and-outbreaks-occupational-settings-eueea-and-uk.

47. State of California. Employment Development Department (EDD). Labor market information. Accessed November 8, 2021. https://www.labormarketinfo.edd.ca.gov/.

48. Centers for Disease Control and Prevention. COVID-19: workplaces and businesses. Accessed November 8, 2021. https://www.cdc.gov/coronavirus/2019-ncov/community/workplaces-businesses/index.html.

49. State of California. Division of Occupational Safety and Health (Cal/OSHA). COVID-19 Prevention Emergency Temporary Standards. Accessed November 8, 2021. https://www.dir.ca.gov/dosh/coronavirus/ETS.html.

50. Hanage WP, Testa C, Chen JT, et al. COVID-19: US federal accountability for entry, spread, and inequities-lessons for the future. Eur J Epidemiol. 2020;35(11):995–1006.

51. Carlsten C, Gulati M, Hines S, et al. COVID-19 as an occupational disease. Am J Ind Med. 2021;64(4):227–237.

